# Delayed Interventions, Low Compliance, and Health Disparities Amplified the Early Spread of COVID-19

**DOI:** 10.1101/2020.07.31.20165654

**Authors:** Aliea M. Jalali, Sumaia G. Khoury, JongWon See, Alexis M. Gulsvig, Brent M. Peterson, Richard S. Gunasekera, Gentian Buzi, Jason Wilson, Thushara Galbadage

## Abstract

The United States (US) public health interventions were rigorous and rapid, yet failed to arrest the spread of the Coronavirus Disease 2019 (COVID-19) pandemic as infections spread throughout the US. Many factors have contributed to the spread of COVID-19, and the success of public health interventions depends on the level of community adherence to preventative measures. Public health professionals must also understand regional demographic variation in health disparities and determinants to target interventions more effectively. In this study, a systematic evaluation of three significant interventions employed in the US, and their effectiveness in slowing the early spread of COVID-19 was conducted. Next, community-level compliance with a state-level stay at home orders was assessed to determine COVID-19 spread behavior. Finally, health disparities that may have contributed to the disproportionate acceleration of early COVID-19 spread between certain counties were characterized. The contribution of these factors for the disproportionate spread of the disease was analyzed using both univariate and multivariate statistical analyses. Results of this investigation show that delayed implementation of public health interventions, a low level of compliance with the stay at home orders, in conjunction with health disparities, significantly contributed to the early spread of the COVID-19 pandemic.

## 1. Introduction

Amidst the unprecedented early public health response to the Coronavirus Disease 2019 (COVID-19) pandemic, the United States (US) was unable to contain the spread of the Coronavirus. Public health officials implemented rigorous evidence-based nonpharmaceutical interventions (NPIs) originating from measures utilized during the 1918 influenza pandemic to help “flatten the curve” (Ferguson et al., 2006; Bootsma and Ferguson, 2007; Hatchett et al., 2007). The effectiveness of NPIs implemented was dependent on four factors that included the concurrent use of NPIs, early intervention, duration, and the rigor of the preventative measure (Hatchett et al., 2007). Seemingly rigorous and early preventative measures that were employed to slow the spread of the COVID-19 have, unfortunately, resulted in subpar and inconsistent outcomes across the US (Jalali et al., 2020).

In the US, public health interventions were mainly administered at the state level. While this caused certain nuances to the nationwide preventative efforts put forth to contain the spread of COVID-19, they were similar in many aspects. Four of the main NPIs implemented in the US included restrictions on mass gatherings, stay at home orders, face mask requirements, and screening for COVID-19. While similar interventions were used in many states, we observed a disproportionate early spread of the Coronavirus in some states, including New York, California, Florida, and Texas (Jalali et al., 2020).

Several factors may have contributed to the observed differences in the propagation of the pandemic. These fall into three categories. (1) The effectiveness and the timeliness of the public health interventions applied. (2) Community compliance and adherence to the implemented preventative measures. (3) Underlying health disparities unequally distributed across different geographic locations. However, to our knowledge, no formal determination as to how these factors collectively contributed to the uneven spread of the COVID-19 pandemic has been made. Specifically, no systematic evaluation of public health interventions on the slowing or stopping of the COVID-19 spread has been conducted.

In a study that characterized the implementation of NPIs in the US during the early stages of the pandemic, the author described preventative measures that were likely to help mitigate the spread of the Coronavirus (Schuchat, 2020). Recently, we compared the implementation of nationwide stay at home orders and evaluated the effectiveness in reducing the COVID-19 case rates in four states (Jalali et al., 2020). Besides the rigor of the interventions employed, it is crucial to identify whether they are followed at the community level. The level of compliance will factor in determining the efficacy of interventions utilized. This aspect of NPIs has also not been well characterized.

Recent studies have focused on the role of health disparities and the social determinants of health in the spread and mortality of this disease in the US (Holmes et al., 2020; Wilder, 2020). Race and ethnicity are factors that many researchers have identified during the COVID-19 pandemic. Investigators studying COVID-19 cases within a Louisiana hospital found that 76.9% of the patients hospitalized with COVID-19 and 70.6% of those who died were African American, although African Americans comprised only 31.0% of the sample (Price-Haywood et al., 2020). Another study in California showed African American patients were 2.7 times more likely to be hospitalized for COVID-19 than Caucasian patients (Azar et al., 2020). Besides racial and ethnic health disparities, the social determinants of health, including socioeconomic status (SES), were highlighted in recent studies. These factors can sometimes be interconnected. African Americans reportedly have greater exposure to environmental pollutants, food insecurity, poor living conditions, illiteracy, low SES, and a lack of healthcare resources, making them more susceptible to COVID-19 fatality (Holmes et al., 2020). Minority groups, including African Americans, Hispanics, and Native Americans, are reported to be more likely to experience socioeconomic disadvantages at some point in their life than non-minority groups (Williams et al., 2010). In addition, previous studies have shown that low-income groups are at an increased risk for mental illness, chronic diseases, lower life expectancy, and higher mortality (Belle Doucet, 2003; Braveman et al., 2010; Mode et al., 2016). The Center for Disease Control and Prevention (CDC) has stated that underlying health conditions and comorbid conditions are major risk factors for COVID-19, thereby subjecting individuals from minority populations to an even greater risk (CDC, 2020).

When addressing the spread of COVID-19, it is important to take into account many influencing factors that may lead to the disproportionate spread of the disease. These may include, but not be limited to, the effectiveness of the preventative measure, community-level compliance to stay at home orders, and underlying health disparities. In this study, we characterized these three aspects in detail as we evaluated the early spread of COVID-19 in the US. We compared these factors individually and collectively in the 30 most populous US counties, to identify possible associations with the disproportionate spread of the disease.

## 2. Materials and Methods

### 2.1 COVID-19 case rates, mortality rates, and case-fatality rates

The COVID-19 county-level number of cases and deaths were obtained from the data provided by the Johns Hopkins University COVID-19 data repository (JHU, 2020). Case rates, mortality rates, and case-fatality rates were calculated from this dataset. The COVID-19 case rates were defined as the cumulative number of cases per unit county population on a given date. Mortality rates were defined as the cumulative number of deaths per unit county population on a given date. Case-fatality rates were defined as the cumulative number of deaths per unit cases in a county on a given date. Case rates, mortality rates, and case-fatality rates were studied from March 1^st^ to May 31^st^, 2020, and were followed for 92 days after each county reached a case rate of 1 per 10,000 and a mortality rate of 1 per 10,000, respectively.

### 2.2 Study population, US county, and spread group selection

To characterize the early COVID-19 spread across the US, we selected the 30 most populous counties in the US and calculated their case rates, mortality rates, and case-fatality rates. These counties were divided into three groups (high, mid, and low) based on their case rates on May 10^th^, 2020. Ten counties were placed in each group according to their case rates. The high group included case rates of > 100 per 10,000, the ten counties in the mid group had case rates of 15 to 100 per 10,000, and the ten counties in the low group had case rates < 15 per 10,000 (Table 1, Supplemental Figure 1). We then characterized these counties’ public health interventions, level of compliance to these interventions, and health disparities to identify factors that influenced the differential spread of COVID-19 in the US. Next, we looked at the ten most populous counties in four states, California, Florida, New York, and Texas, to identify and confirm factors that caused the disproportionate spread of COVID-19 within these states.

**Table 1.**
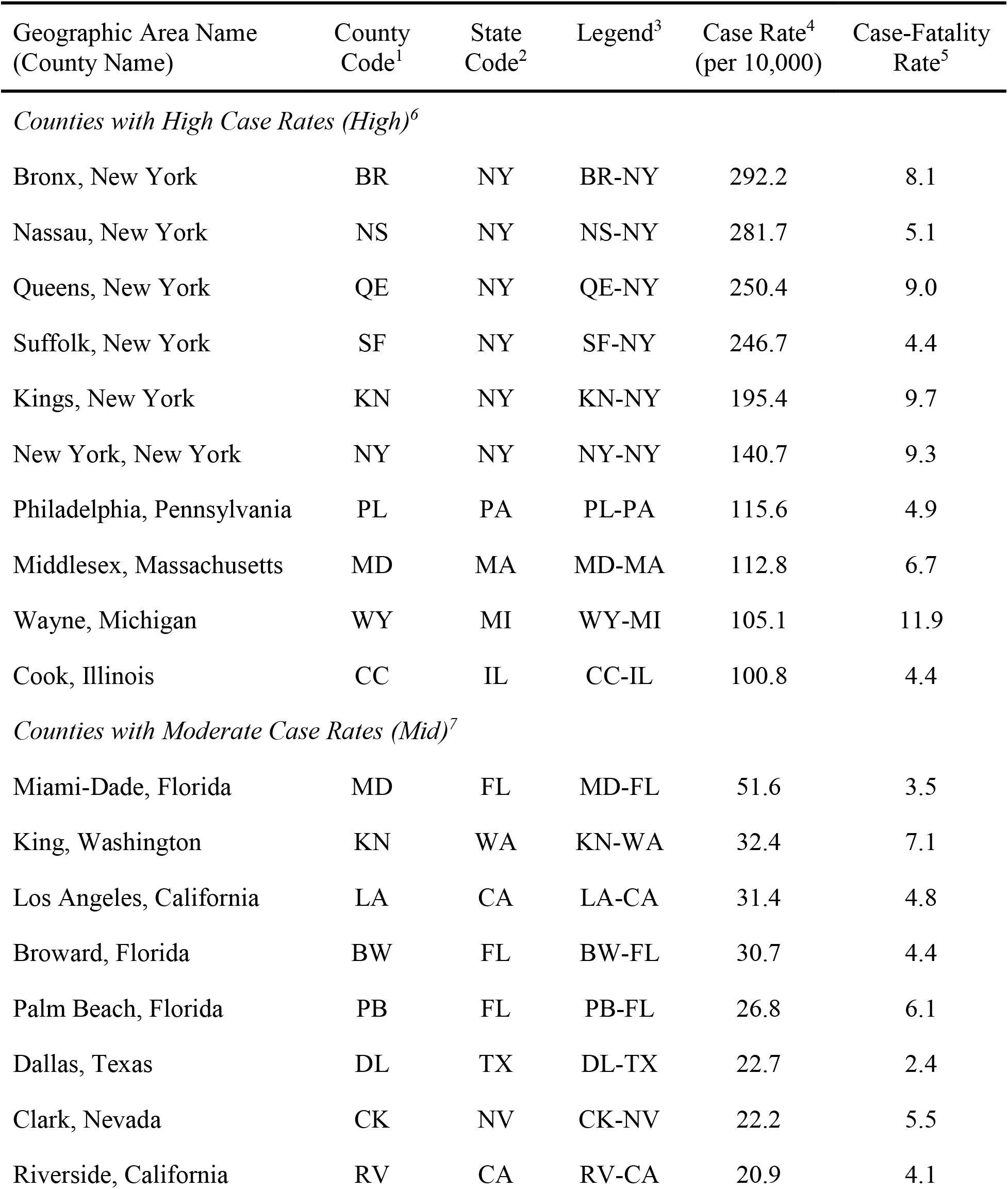
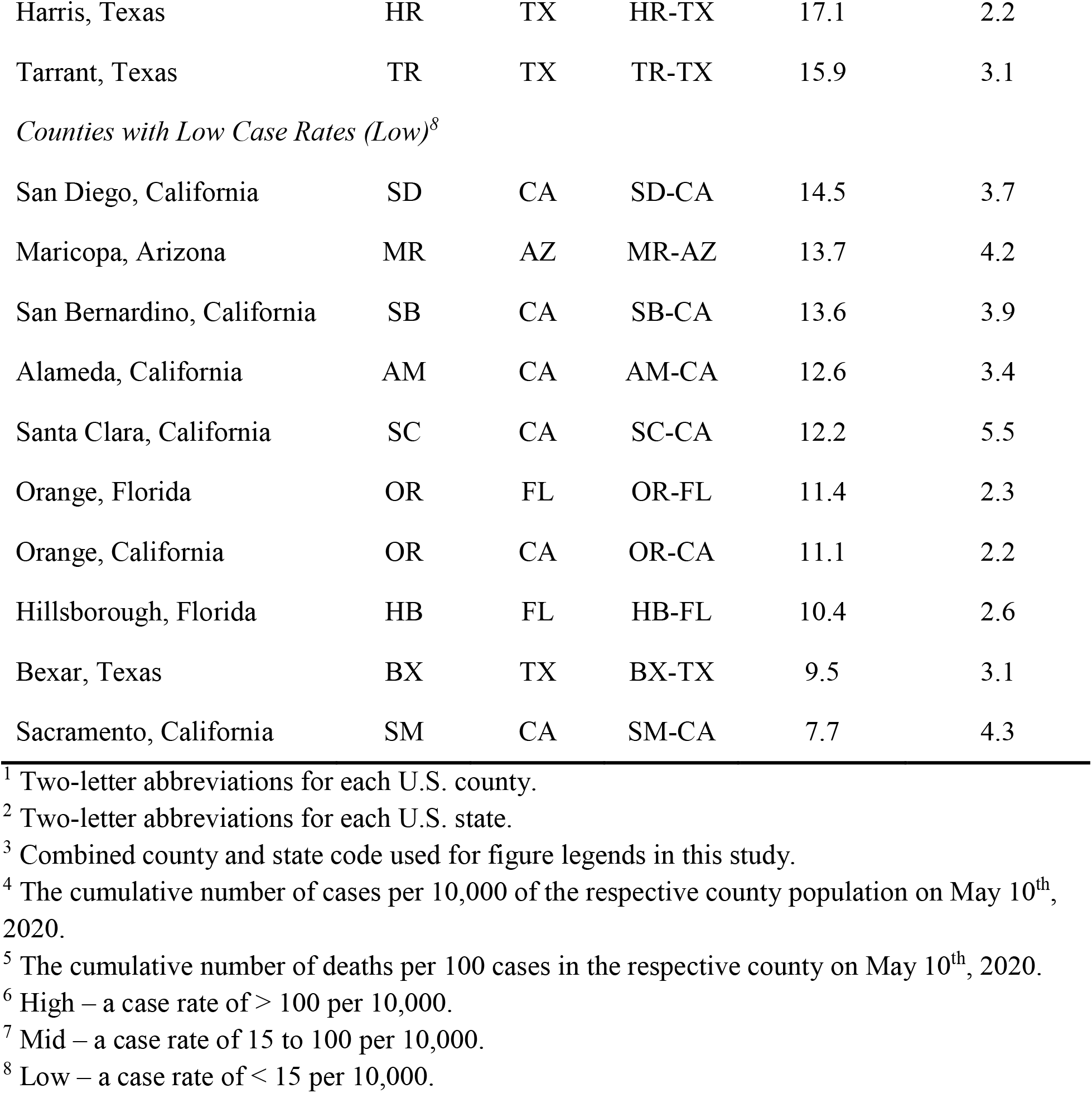
COVID-19 Case rates and case-fatality rates of the 30 most populous counties in the United States on May 10^th^, 2020.

| Geographic Area Name<br>(County Name) | County<br>Code<sup>1</sup> | State<br>Code<sup>2</sup> | Legend<sup>3</sup> | Case Rate<sup>4</sup><br>(per 10,000) | Case-Fatality<br>Rate<sup>5</sup> |
| --- | --- | --- | --- | --- | --- |
| *Counties with High Case Rates (High)<sup>6</sup>* |  |  |  |  |  |
| Bronx, New York | BR | NY | BR-NY | 292.2 | 8.1 |
| Nassau, New York | NS | NY | NS-NY | 281.7 | 5.1 |
| Queens, New York | QE | NY | QE-NY | 250.4 | 9.0 |
| Suffolk, New York | SF | NY | SF-NY | 246.7 | 4.4 |
| Kings, New York | KN | NY | KN-NY | 195.4 | 9.7 |
| New York, New York | NY | NY | NY-NY | 140.7 | 9.3 |
| Philadelphia, Pennsylvania | PL | PA | PL-PA | 115.6 | 4.9 |
| Middlesex, Massachusetts | MD | MA | MD-MA | 112.8 | 6.7 |
| Wayne, Michigan | WY | MI | WY-MI | 105.1 | 11.9 |
| Cook, Illinois | CC | IL | CC-IL | 100.8 | 4.4 |
| *Counties with Moderate Case Rates (Mid)<sup>7</sup>* |  |  |  |  |  |
| Miami-Dade, Florida | MD | FL | MD-FL | 51.6 | 3.5 |
| King, Washington | KN | WA | KN-WA | 32.4 | 7.1 |
| Los Angeles, California | LA | CA | LA-CA | 31.4 | 4.8 |
| Broward, Florida | BW | FL | BW-FL | 30.7 | 4.4 |
| Palm Beach, Florida | PB | FL | PB-FL | 26.8 | 6.1 |
| Dallas, Texas | DL | TX | DL-TX | 22.7 | 2.4 |
| Clark, Nevada | CK | NV | CK-NV | 22.2 | 5.5 |
| Riverside, California | RV | CA | RV-CA | 20.9 | 4.1 |
| Harris, Texas | HR | TX | HR-TX | 17.1 | 2.2 |
| Tarrant, Texas | TR | TX | TR-TX | 15.9 | 3.1 |
| *Counties with Low Case Rates (Low)<sup>8</sup>* |  |  |  |  |  |
| San Diego, California | SD | CA | SD-CA | 14.5 | 3.7 |
| Maricopa, Arizona | MR | AZ | MR-AZ | 13.7 | 4.2 |
| San Bernardino, California | SB | CA | SB-CA | 13.6 | 3.9 |
| Alameda, California | AM | CA | AM-CA | 12.6 | 3.4 |
| Santa Clara, California | SC | CA | SC-CA | 12.2 | 5.5 |
| Orange, Florida | OR | FL | OR-FL | 11.4 | 2.3 |
| Orange, California | OR | CA | OR-CA | 11.1 | 2.2 |
| Hillsborough, Florida | HB | FL | HB-FL | 10.4 | 2.6 |
| Bexar, Texas | BX | TX | BX-TX | 9.5 | 3.1 |
| Sacramento, California | SM | CA | SM-CA | 7.7 | 4.3 |

<sup>1</sup> Two-letter abbreviations for each U.S. county.
<sup>2</sup> Two-letter abbreviations for each U.S. state.
<sup>3</sup> Combined county and state code used for figure legends in this study.
<sup>4</sup> The cumulative number of cases per 10,000 of the respective county population on May 10<sup>th</sup>, 2020.
<sup>5</sup> The cumulative number of deaths per 100 cases in the respective county on May 10<sup>th</sup>, 2020.
<sup>6</sup> High – a case rate of > 100 per 10,000.
<sup>7</sup> Mid – a case rate of 15 to 100 per 10,000.
<sup>8</sup> Low – a case rate of < 15 per 10,000.

### 2.3 State and county-level public health interventions

We surveyed the public health interventions and actions implemented at the state and county-level from January 1^st^ to July 1^st,^ 2020 (Supplemental Table 1-6). These data were extracted from the state and county public health department and government websites. We examined press releases, executive orders issued by each state, and their counties to determine various public health interventions enacted. Using this information, we organized public health interventions into three broad categories. They were (1) restrictions on mass gatherings (Supplemental Table 1 and 2), (2) stay at home orders (Supplemental Table 3 and 4), and (3) face mask requirements (Supplemental Figure 5 and 6). For each of the counties in this study, we characterized the quality and intensity of their public health interventions until May 10^th^, 2020, focusing on the following four specific criteria. (1) The duration of the corresponding intervention was implemented. (2) The number of days the corresponding intervention was delayed before it started and a case rate of 1 per 10,000 was used as a reference start date. (3) The number of COVID-19 cases in each county the day before the start of the corresponding intervention. (4) The COVID-19 case rate in each county, the day before the start of the corresponding public health response. The public health interventions were then compared in the three COVID-19 spread groups (high, mid, and low).

### 2.4 Assessing community compliance with public health interventions

We used Google COVID-19 Community Mobility Reports and the Unacast Social Distancing Scoreboard to assess the publics’ compliance with public health interventions enacted by their state or county (Google, 2020; Unacast, 2020). The Google dataset provided mobility trends showing percent change in the number of visits over time by geography, across different categories of places. The baseline was six weeks of pre-COVID-19 (before March 2020) using anonymously collected google location history data. Location included (a) retail and recreation, (b) groceries and pharmacies, (c) parks, (d) transit stations, (e) workplaces, and (f) residential. Percent change in the residential category represented a change in duration while all other categories represented a change in the total number of visitors. The Unacast dataset provided mobile device location data. Devices were assigned to counties based on where a specific device was recorded for the longest time on a particular day. The pre-COVID-19 period was defined as four weeks before March 8^th^, 2020. Percent changes in the movement are shown in three categories: (g) distance traveled, (h) non-essential points of interest (POIs) visitation, and (i) human encounters. Total overall movement (j) includes a combined score for the distance traveled, POIs visitation, and human encounters. Change in mobility and movement was calculated as the mean of daily percent changes from the start of stay at home orders until May 10^th^, 2020, or the end of the stay at home orders (whichever came first), for the corresponding county. For each of the above categories (a-j), we plotted the time series data for percent change of mobility as a 7-day rolling average for the high, mid, and low groups.

### 2.5 Characterizing health disparities across selected counties

Demographic and social determinants of health data from the 2018 US Census with five-year estimates (when available) were obtained for all the counties in this current study (USCB, 2020). For age and sex data, table S0101 “Age and Sex” were used. Age ranges studied included years 0-19, 20-44, 45-64, >65. We compared the percent population that was ≥ 65 years old in each of the counties. The land area of each county was obtained from table LND01 2011 dataset. The population density was calculated using total county population information and land area for each county in square miles. For race and ethnicity, we used table B03002 “Race and Ethnicity.” We compared the percent population that was Caucasian, Hispanic, African American, and Asian. For income and poverty data, table B17002 “Income and Poverty” were used. Only a 1-year estimate was available for the B17002 dataset. This table provides data on the ratio of income to the poverty level in the past twelve months. For this study, we used ratio ranges for < 0.99 (below poverty line) and > 5 times the poverty level (higher income). For data on housing units, table B25001 “Housing Units” was used, and information regarding housing density was collected. Table B25008, “Rent or Own Houses,” was used to gather data on the percent population that rents or owns their homes. For information on household and family size table S1101, “Households and Families” was used. Table B27001, “Health Insurance,” was used to determine the percentage of individuals without health insurance. To determine the level of education attainment table B15003, “Education Attainment 25 years and older,” was used. We used the percent population that had at least one year of college education in this study. We used the Johns Hopkins University COVID-19 Resource Center to collect state information on the number of staffed hospital beds and intensive care unit (ICU) beds available in each of the study counties (JHU, 2020). We compared these health determinants among the three COVID-19 spread groups (high, mid, and low) to identify any health disparities that may have contributed to the disproportionate spread of the Coronavirus in the US.

### 2.6 Statistical Analysis

Non-parametric Kruskal-Wallis tests were employed to compare the three COVID-19 spread groups (high, mid, and low) in individual categories. These analyses characterized the associations between the differential spread of COVID-19 in the US and various parameters, including public health interventions, community compliance, and health disparities. Pearson’s correlation coefficients and partial correlation coefficients were calculated for all pairs of numeric variables under study. We also performed multiple regression analyses for each of the three categories, (1) public health interventions, (2) community compliance, and (3) health determinants to assess the combined contribution of each of these parameters.

## 3. Results

### 3.1 US most populous counties displayed disproportionate COVID-19 case rates, mortality rates, and case-fatality rates

To show that there were differences in the early spread of the COVID-19 pandemic across the US, we examined COVID-19 case rates in the 30 most populous US counties. On May 10^th^, 2020, we observed case rates of 7.7 to 292.2 per 10,000 county population, even among these most populous US counties (Table 1, Supplemental Table 1). We grouped these counties into high, mid, and low case rate groups, with ten counties in each spread group (Table 1). We then plotted case rates for these counties from March 1^st^ to May 31^st^, 2020, and the mean case rates for the high, mid, and low spread groups (Figure 1 a-d). We also looked at case rates in two-week intervals from April 12^th^ to May 24^th^, 2020, and noted significant differences between these three groups [p < 0.0001] (Figure 1 e-h). While these were the most populous US counties and were affected very early in the pandemic, the start of the COVID-19 spread had some temporal differences. To account for the delay in the increase of cases among these counties, we used 1 case per 10,000 as a point of reference and followed the increase in case rates for the next 92 days (13 weeks) (Figure 1 i-l). The increase in the case rates showed different patterns among the three groups. The high spread group showed an initial exponential increase followed by a plateauing of their case rates. The mid group had a somewhat constant increase in case rates, while the low group displayed a slow rise in cases initially followed by a delayed exponential rise in their case rates (Figure 1 i-k). Fourteen-day (2 weeks) intervals showed that there were consistently significant differences in the case rates among these three groups temporally. [p < 0.0001] (Figure 1 m-p). These results showed that there were significant differences in the spread of the early pandemic in the 30 most populous US counties.

Next, we evaluated the mortality rates for the counties in the high, mid, and low spread groups similar to our analyses for case rates. We found that higher spread groups had the highest mortality rates, followed by the mid spread group and the low spread groups (Supplemental Figure 2). To examine if there were differences in the number of deaths per case in the high, mid, and low spread groups, we calculated the case-fatality rates in each of the counties on May 10^th^, 2020 (Table 1). Case-fatality rates ranging from 2.2 to 11.9 per 100 cases were observed among these counties. To evaluate the differences in these case-fatality rates, we plotted case-fatality rates for these counties from March 1^st^ to May 31^st^, 2020 (Figure 2 a-c). The mean case-fatality rates for the high spread group were significantly greater than the case-fatality rates in the mid and low spread groups (Figure 2 d). To characterize these differences further, we looked at case-fatality rates in two-week intervals from April 12^th^ to May 24^th^, 2020. We noted a significant difference among these three groups at all-time points [p = 0.0412, p = 0.0117, p = 0.0011, and p = 0.0006 respectively] (Figure 2 e-h). Among all 30 counties, the correlation between case rate and the mortality rate was 0.93, between case rates and case-fatality rates was 0.57, and between mortality rate and case-fatality rate was 0.76.

**Figure 1.**
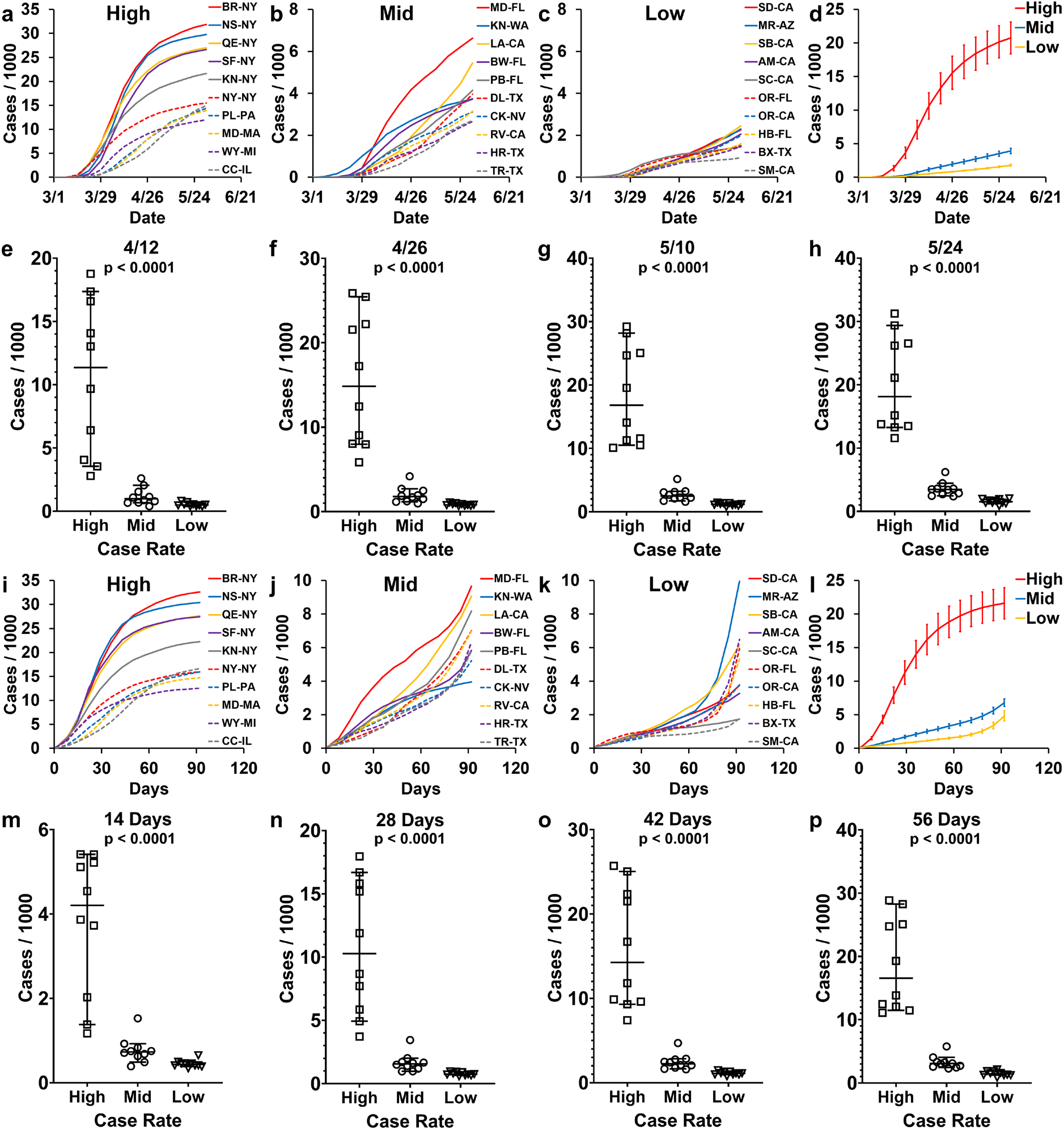
COVID-19 case rates in the 30 most populous US counties. COVID-19 case rates defined as the cumulative number of cases per unit county population. The 30 most populous US counties divided into three groups (high, mid, and low) based on their COVID-19 case rates on May 10^th^, 2020. The high group with case rates of > 100 per 10,000, the mid group with case rates of 15 to 100 per 10,000 and low group with case rates < 15 per 10,000. The counties included in the high, mid, and low groups are listed in Table 1 with the figure legend abbreviations. (a-h) COVID-19 cases rates from March 1^st^, 2020 to May 31^st^, 2020, in the respective county. (a) The case rates of counties with high case rates. (b) The case rates of counties with moderate (mid) case rates. (c) The case rates of counties with low case rates. (d) The mean case rates of counties in high, mid, and low groups with error bars indicating the standard error. (e-h) Cumulative cases per 1000 in each of the three groups on a specific date. Median and 95% confidence interval (95% CI) are presented. (e) April 12^th^, 2020. (f) April 26^th^, 2020. (g) May 10^th^, 2020. (h) May 24^th^, 2020. (i-p) COVID-19 cases rates in each county, starting at a case rate of 1 per 10,000, followed for the next 92 days. (i) The case rates of counties with high case rates. (j) The case rates of counties with moderate (mid) case rates. (k) The case rates of counties with low case rates. (l) The mean case rates of counties in high, mid, and low groups with error bars indicating the standard error. (m-p) Cumulative cases per 1000 in each of the three groups on a specific day, starting at a case rate of 1 per 10,000. Median and 95% confidence interval (95% CI) are presented. (m) Fourteen (14) days post reaching a case rate of 1 per 10,000. (n) Twenty-eight (28) days post reaching a case rate of 1 per 10,000. (g) Forty-two (42) days post reaching a case rate of 1 per 10,000. (h) Fifty-six (56) days post reaching a case rate of 1 per 10,000. A non-parametric Kruskal-Wallis test was performed to compare the three groups (high, mid, and low), and the p-values are indicated above each of the corresponding results.

**Figure 2.**
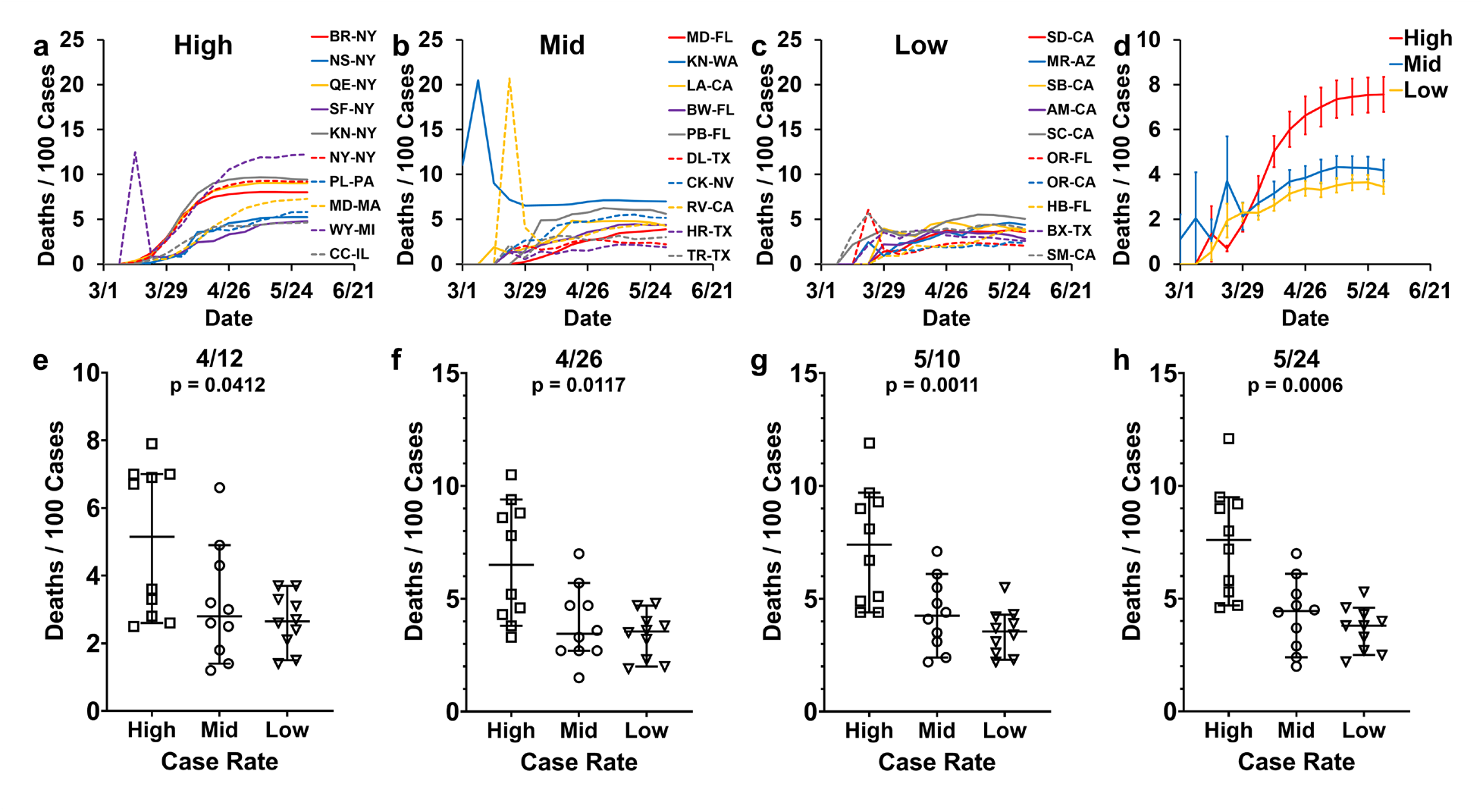
COVID-19 case-fatality rates in the 30 most populous US counties. COVID-19 case-fatality rates defined as the cumulative number of deaths per unit cases in the county. The 30 most populous US counties divided into three groups (high, mid, and low) based on their COVID-19 case rates on May 10^th^, 2020. The counties included in the high, mid, and low groups are listed in Table 1 with the figure legend abbreviations. (a-h) COVID-19 case-fatality rates from March 1^st^, 2020 to May 31^st^, 2020, in the respective county. (a) The case-fatality rates of counties with high case rates. (b) The case-fatality rates of counties with moderate (mid) case rates. (c) The case-fatality rates of counties with low case rates. (d) The mean case-fatality rates of counties in high, mid, and low groups with error bars indicating the standard error. (e-h) Cumulative deaths per 100 cases in each of the three groups on a specific date. Median and 95% confidence interval (95% CI) are presented. (e) April 12^th^, 2020. (f) April 26^th^, 2020. (g) May 10^th^, 2020. (h) May 24^th^, 2020. A non-parametric Kruskal-Wallis test was performed to compare the three groups (high, mid, and low), and the p-values are indicated above each of the corresponding results.

### 3.2 Early public health interventions helped slow the spread of COVID-19

Assessment of the three main interventions implemented across the US at both the state and county levels, including restrictions on mass gatherings, stay at home orders, and facemask requirements, were conducted. This included the duration of the intervention, delay in implementation, number of cases, and the case rate at the start of these actions within the high, mid, and low case rate groups (Figure 3). Results demonstrated that restrictions on mass gatherings alone were not associated with changes in the early spread of COVID-19 (Figure 3 a-d). There were no statistically significant differences between the high, mid, and low spread counties for duration [p = 0.20306], delay in implementation [p = 0.2346], number of cases [p = 0.2022], or the case rate [p = 0.3826] for restrictions on mass gatherings (Figure 4 a-d).

For stay at home orders alone there was a significant difference between high, mid, and low spread counties for duration [p = 0.0219], delay in implementation [p = 0.0206], number of cases [p = 0.0023], and case rate [p = 0.0036] (Figure 3 e-h). Stay at home orders had longer durations in the high spread counties compared to mid and low spread counties (Figure 3 e). The longer durations could reflect counties with high case burdens implementing more rigorous measures. The differences in delay in enacting stay at home orders, number of cases and case rate at the start of the intervention suggests that the early implementation of stay at home orders was associated with lower case rates of COVID-19 (Figure 3 f-h). Duration and delay in the implementation of face mask requirements alone showed no significant difference between the high, mid, and low spread groups [p = 0.3395 and p = 0.2339, respectively] (Figure 3 i,j). The bimodal distribution observed in the duration and delay of the low spread group may, in part, account for this observation. There was a significant difference among the three spread groups for the number of cases and the case rate at the time of implementation of face mask requirements [p = 0.0001 and p < 0.0001 respectively] (Figure 3 k,l). This difference shows that the early introduction of face mask requirements alone was associated with significantly lower case rates.

**Figure 3.**
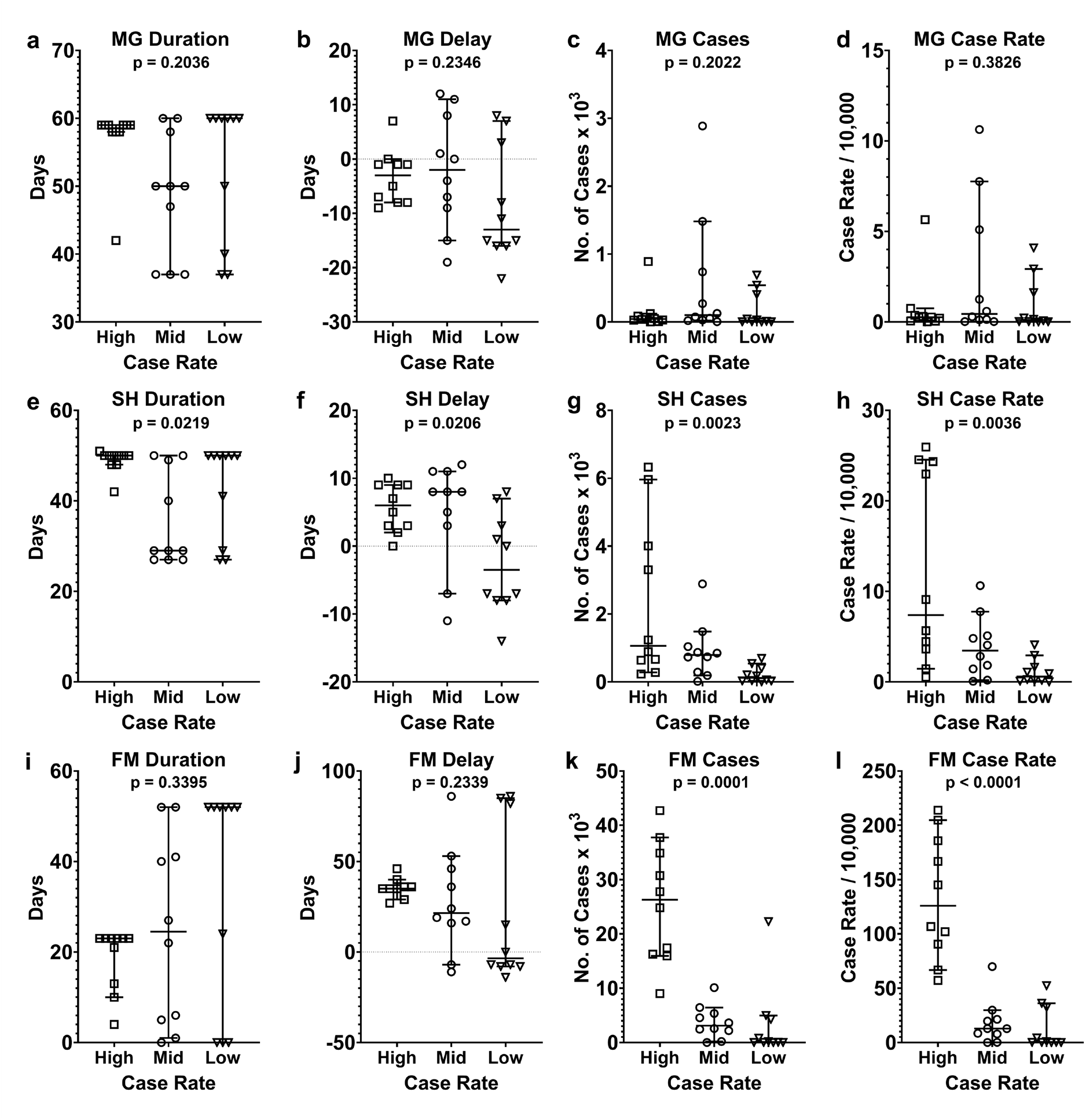
Characterization of public health response in the 30 most populous counties in the US. The 30 most populous US counties divided into three groups (high, mid, and low) based on their COVID-19 case rates on May 10th, 2020. (a-d) Restrictions on mass gatherings (MG). (e-h) Stay at home orders (SH). (i-l) Face mask (FM) mandate. Median and 95% confidence interval (95% CI) are presented. (a, e, i) The duration of the corresponding intervention was implemented. (b, f, j) The number of days the corresponding intervention was delayed before it started. A case rate of 1 per 10,000 was used as a reference start date. (c, g, k) The number of COVID-19 cases in each county the day before the start of the corresponding intervention. (d, h, l) The COVID-19 case rate in each county, the day before the start of the corresponding intervention. A non-parametric Kruskal-Wallis test was performed to compare the three groups (high, mid, and low), and the p-values are indicated above each of the corresponding results.

### 3.3 Low compliance caused COVID-19 to spread amidst rigorous interventions

Rigorous public health interventions can only be effective to the degree to which the intended population follows them. Google community mobility reports were utilized to determine if the state and county level public health measures were effectively implemented (Figure 4 a-f). The percent change in mobility in the high, mid, and low spread counties was evaluated to determine the level of compliance with the stay at home orders implemented. We defined an 81 to 100% reduction in mobility as high compliance, a 51 to 80% reduction mobility as moderate compliance, and any present change ≤ 50% as low compliance. We observed the following median reductions of mobility in each group, compared to a pre-COVID-19 baseline. Retail and recreation locations: high 55.2%, mid 46.0%, and low 47.4% [p = 0.026] (Figure 4 a). Grocery and pharmacies: high 20.0%, mid 17.3%, and low 18.1% [p = 0.4036] (Figure 4 b). Parks: high 11.3%, mid 43.7%, and low 32.0% [p = 0.1344] (Figure 4 c). Transit stations: high 63.8%, mid 57.4%, and low 57.0% [p = 0.3476] (Figure 4 d). Workplaces: high 56.5%, mid 46.5%, and low 48.1% [p = 0.005] (Figure 4 e). Residential locations showed an increase: high 24.2%, mid 19.8%, and low 20.2% [p = 0.0384] (Figure 4 f). A significant difference in mobility was observed between high, mid, and low spread groups in the mobility observed at retail and recreation locations [p = 0.0206], workplaces [p = 0.005], and residential locations [p = 0.0384]. Our results indicate that even during the period when stay at home orders was enforced, mobility reduction occurred sporadically with as little as 0-to-20.0% reductions in visitations of certain locations. While visitations to certain other locations were reduced as much as 55.0 to 75.0%, the underlying non-compliance observed at the community level makes seemingly rigorous public health interventions ineffective.

To confirm our Google community mobility report results, we compared mobile device location data provided by Unacast, in our study counties (Unacast, 2020) (Figure 4 g-j). Compared to a pre-COVID-19 baseline, the following percentage decreases in the movement were observed. Distance traveled: high 51.6%, mid 46.5%, and low 45.9% [p = 0.1566] (Figure 4 g). Non-essential POI visitations: high 67.6%, mid 63.0%, and low 64.4% [p = 0.1946] (Figure 4 h). Human encounters: high 35.0%, mid 35.0% and low 35.2% [p = 0.1556] (Figure 4 i). Total overall movement: high 59.4%, mid 52.6%, and low 54.4% [p = 0.2125] (Figure 4 j). Unacast mobility data showed no significant difference in the reduction of movement among the high, mid, and low spread groups. Our analyses showed a reduction in movements of 20-to-70.0% across different parameter highlights that there was low compliance to the stay at home orders enforced and supports our conclusions from the Google mobility reports.

**Figure 4.**
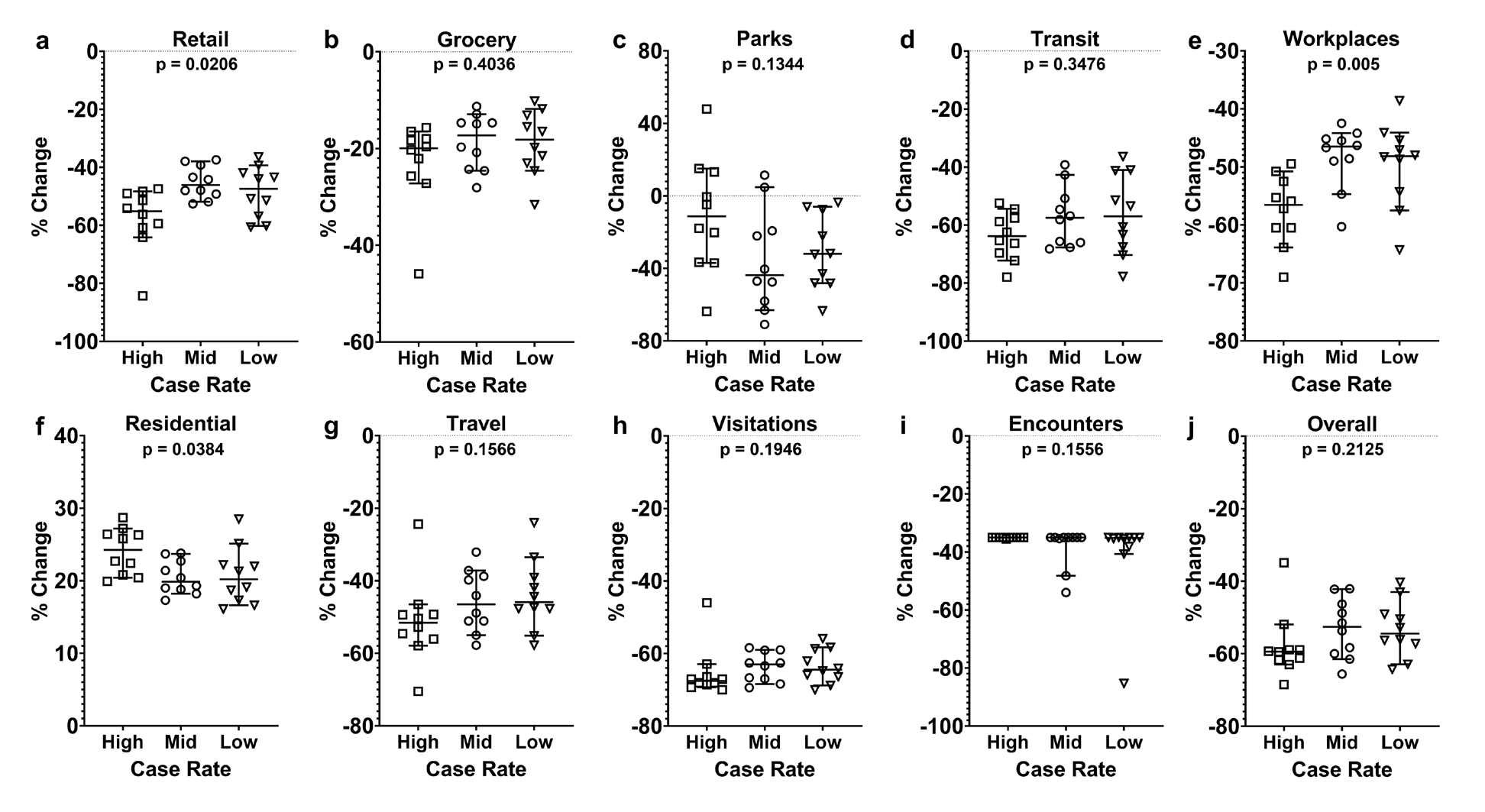
Change in community mobility during the period stay at home orders were implemented. Change in mobility and movement shown as the mean of daily percent changes from the start of stay at home orders until May 10^th^, 2020, or the end of the stay at home orders (whichever came first), for the corresponding county. Median and 95% confidence interval (95% CI) are presented. The 30 most populous US counties divided into three groups (high, mid, and low) based on their COVID-19 case rates on May 10th, 2020. High group with case rates of > 100 per 10,000, mid group with case rates of 15 to 100 per 10,000, and low group with case rates < 15 per 10,000. (a-f) Google community mobility trends showing percent change in movement over time by geography, across different categories of places. The baseline was six weeks of pre-COVID-19 (before March 2020) using anonymously collected google location history data (Google, 2020). Location included (a) retail and recreation, (b) groceries and pharmacies, (c) parks, (d) transit stations, (e) workplaces, and (f) residential. Percent change in Residential was calculated by the duration, while the other categories (a-e) were by the number of visits. (g-j) Unacast mobile device location data (Unacast, 2020). Devices were assigned to counties based on where a specific device was recorded for the longest time on a particular day. The pre-COVID-19 period was defined as four weeks before March 8^th^, 2020. Percent changes in the movement are shown in four categories, (g) distance traveled, (H) non-essential points of interest (POIs) visitation, and (i) human encounters. (j) Total overall movement includes a combined score for the distance traveled, POIs visitation, and human encounters. A non-parametric Kruskal-Wallis test was performed to compare the three groups (high, mid, and low), and the p-values are indicated above each of the corresponding results.

To further confirm our observations of community-level low compliance with stay at home orders, we used time-series data from both Google mobility reports and Unacast data. We plotted the average change in movements for the high, mid, and low spread groups from March 1^st^ to May 31^st^, 2020 (Figure 5 a-j). Our results show that the categories of retail, grocery, parks, travel, visitations, human encounters, and overall had a low level of compliance almost throughout the stay at home period. This is also compounded by the day and weekend variability of mobility in each of these categories. Parks, in particular, showed an increase in mobility compared to baseline in most high spread counties (Figure 5 c).

**Figure 5.**
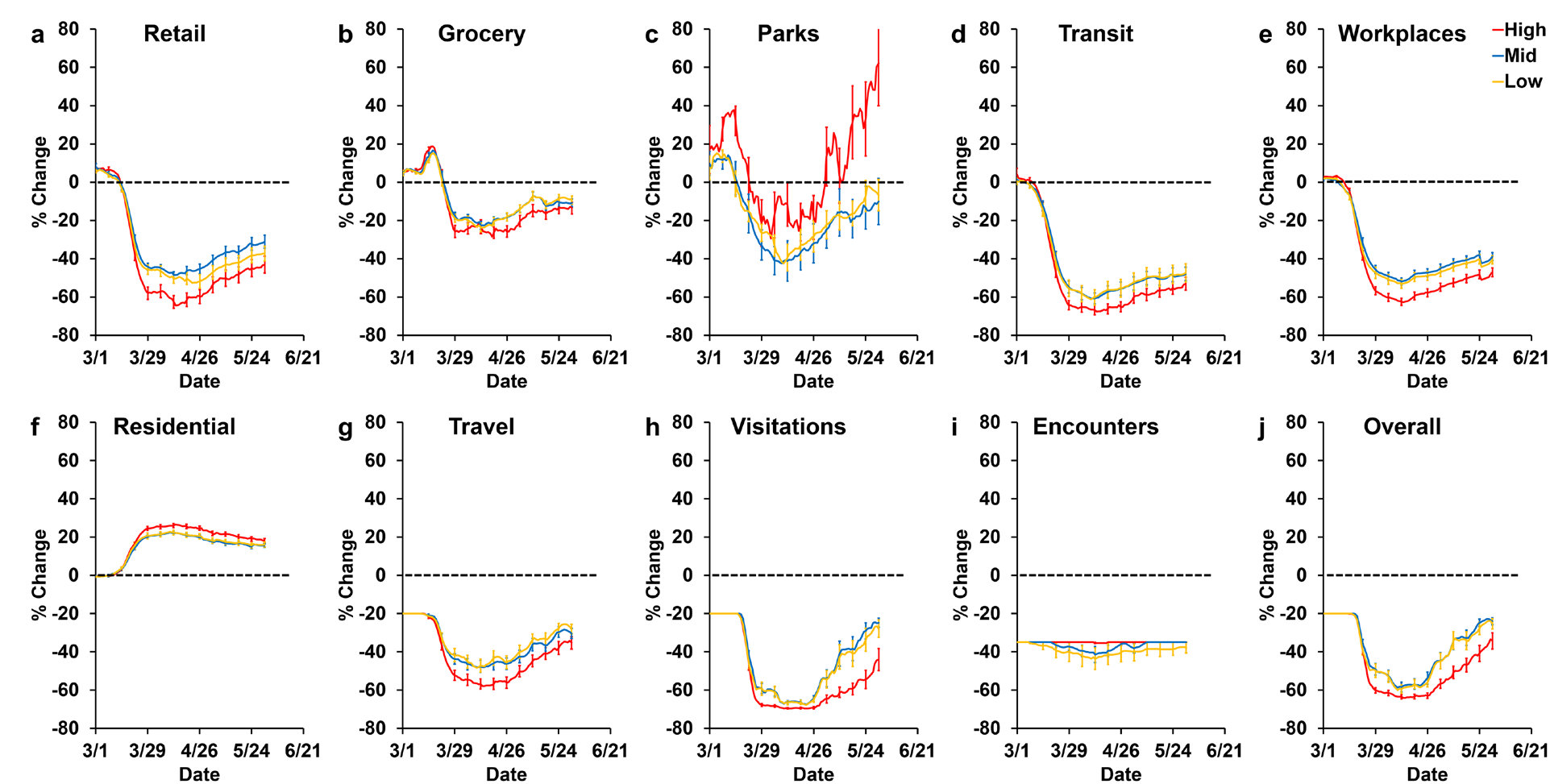
Daily average change in community mobility pre and post COVID-19. Change in mobility and movement shown as the mean of seven day rolling averages of the daily percent changes for each of the COVID-19 spread groups, high, mid and low. Error bars represent the standard error of the mean every seven days. Percent movement changes from March 1^st^ to May 30^th^, 2020. The 30 most populous US counties divided into three groups (high, mid, and low) based on their COVID-19 case rates on May 10th, 2020. High group with case rates of > 100 per 10,000, mid group with case rates of 15 to 100 per 10,000, and low group with case rates < 15 per 10,000. The figure legend is the same for all plots (a-j). The dotted black line represents 0% change in movement pre and post COVID-19. (a-f) Google community mobility trends showing percent change in movement over time by geography, across different categories of places (Google, 2020). Location included (a) retail and recreation, (b) groceries and pharmacies, (c) parks, (d) transit stations, (e) workplaces, and (f) residential. Percent change in Residential was calculated by the duration, while the other categories (a-e) were by the number of visits. (g-j) Unacast mobile device location data (Unacast, 2020). Percent changes in the movement are shown in four categories, (g) distance traveled, (H) non-essential points of interest (POIs) visitation, and (i) human encounters. (j) Total overall movement includes a combined score for the distance traveled, POIs visitation, and human encounters.

### 3.4 Health disparities contribute to the disproportionate spread of COVID-19

A third factor that can contribute to the differences we observe in COVID-19 case rates among the most populous US counties are underlying health disparities. We used US census data to characterize demographic differences, including social determinants of health, to examine associations with the early spread of COVID-19 in the US. We categorized these data into location-based differences (Figure 6 a-e), person-specific characteristics (Figure 6 f-j), and social determinants of health (Figure 6 k-o). These factors were categorically compared to the high, mid, and low spread groups. We observed significant differences among the high, mid, and low spread groups and the following factors when compared individually. Population density [p = 0.0043] (Figure 6 b); land area [p = 0.0025] (Figure 6 c); housing density [p = 0.0026] (Figure 6 d); percent males population [p = 0.0008] (Figure 6 g); percent Hispanic population [p = 0.0196] (Figure 6 i); percent African American population [p = 0.0192] (Figure 6 j); and percent uninsured population [p = 0.0043] (Figure 6 n).

**Figure 6.**
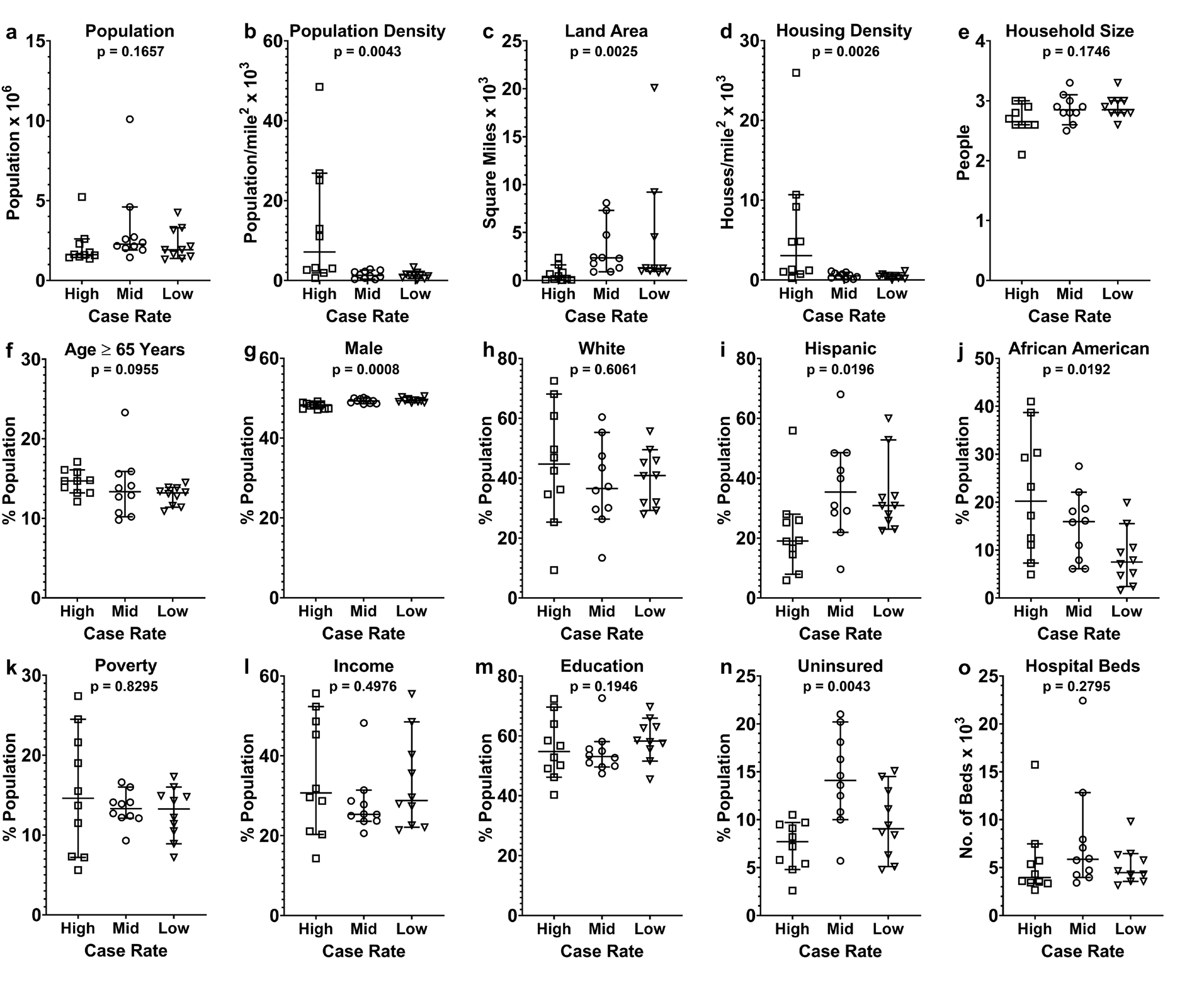
Survey of health determinants and demographic factors that may contribute to the disproportionate COVID-19 spread. Demographic data were obtained from the 2018 US Census data with five-year estimates. The 30 most populous US counties divided into three groups (high, mid, and low) based on their COVID-19 case rates on May 10th, 2020. In each of the groups high, mid and low, case rates are presented as the median and 95% confidence interval (95% CI). (a) Total population in the county. (b) Population density defined as the population per square mile of land area of the county. (c) Land area of the county. (d) Housing density defined as the number of housing units per square mile. (e) Household size defined as the average number of individuals living in a housing unit. (f) Percent population age ≥ 65 years. (g) Percent population male sex. (h) Percent population White ethnicity. (i) Percent population Hispanic ethnicity. (j) Percent population African American Race. (k) Percent population below the poverty line. (l) Percent population with incomes ≥ 5 times the poverty line. (m) Percent population with at least one year of college education. (n) Percent uninsured population. (o) Number of staffed hospital beds in the county. A non-parametric Kruskal-Wallis test was performed to compare the three groups (high, mid, and low), and the p-values are indicated above each of the corresponding results.

These observations show that the early spread of COVID-19 was associated with high population density and housing densities. Given the primary mode of transmission of COVID-19, this is a plausible association (Galbadage et al., 2020). Among our study counties, there was a lower percentage of Hispanics and a higher percentage of African Americans, the Hispanic population, and a higher percentage of African American population in the high spread counties. We did not observe significant differences among the high, mid, and low spread groups and the following factors when compared individually. Total county population [p = 0.1667] (Figure 6 a); average household size [p = 0.1746] (Figure 6 e), percent population with age ≥ 65 years [p = 0.0955] (Figure 6 f); percent Caucasian population [p = 0.6061] (Figure 6 h); percent population with an income below the poverty line [p = 0.8295] (Figure 6 k); percent population with an income > 5 times the poverty line [p = 0.4976] (Figure 6 l); at least one year of college education [p = 0.1946] (Figure 6 m); and the number of staffed hospital beds [p = 0.2795] (Figure 6 o). In our categorical survey of health determinants, we observed several health disparities, including population, density, housing density, the African American population, and uninsured status as potential factors that contributed to the disproportionate early spread of COVID-19 in the US.

Next, we were interested in determining if county level health disparities observed among the high, mid, and low spread groups had any bearing on the case rates and mortality rates of COVID-19. We used demographically stratified COVID-19 cases and deaths for the five counties making up the New York Borough, to compare to the proportion of each of the county populations in the respective demographic category. When relative ratios of the case rates were compared, we observed that age ≥ 65 years (95% C.I. 1.30-2.091), male sex (95% C.I. 1.05-1.12), and African American race (95% C.I. 1.10-1.83) were all risk factors for the disease (Figure 7 a-e). Within the five Borough counties, Hispanic ethnicity (95% C.I. 0.69-1.26) and Caucasian ethnicity (95% C.I. 0.85-1.22) did not show a significant difference in the proportion of the population with COVID-19 (Figure 7 a, d). A similar trend was observed for mortality rates. Relative ratios for mortalities were, age ≥ 65 years (95% C.I. 4.71-5.83), male sex (95% C.I. 1.24-1.29), and African American race (95% C.I. 1.06-1.78) were all risk factors for the disease (Figure 7 a-e). No significant differences were observed between the five-borough counties with Hispanic ethnicity (95% C.I. 0.74-1.45) and White ethnicity (95% C.I. 0.68-1.32). Next, the association between the level of poverty and COVID-19 spread was evaluated, after which a stepwise increase in the case rate, severe case rate, and mortality rate with the increase in poverty (7 f-h) was observed. These results show that disparities in health determinants, including advanced age, male sex, African American race, and poverty were associated with higher case rates of COVID-19. Next, the association between the level of poverty and COVID-19 spread was evaluated, after which a stepwise increase in the case rate, severe case rate, and mortality rate with the increase in poverty (7 f-h) was observed. These results show that disparities in health determinants, including advanced age, male sex, African American race, and poverty, were associated with higher case rates of COVID-19… Next, the association between the level of poverty and COVID-19 spread was evaluated, after which a stepwise increase in the case rate, severe case rate, and mortality rate with the increase in poverty (7 f-h) was observed. These results show that disparities in health determinants, including advanced age, male sex, African American race, and poverty were associated with higher case rates of COVID-19.

**Figure 7.**
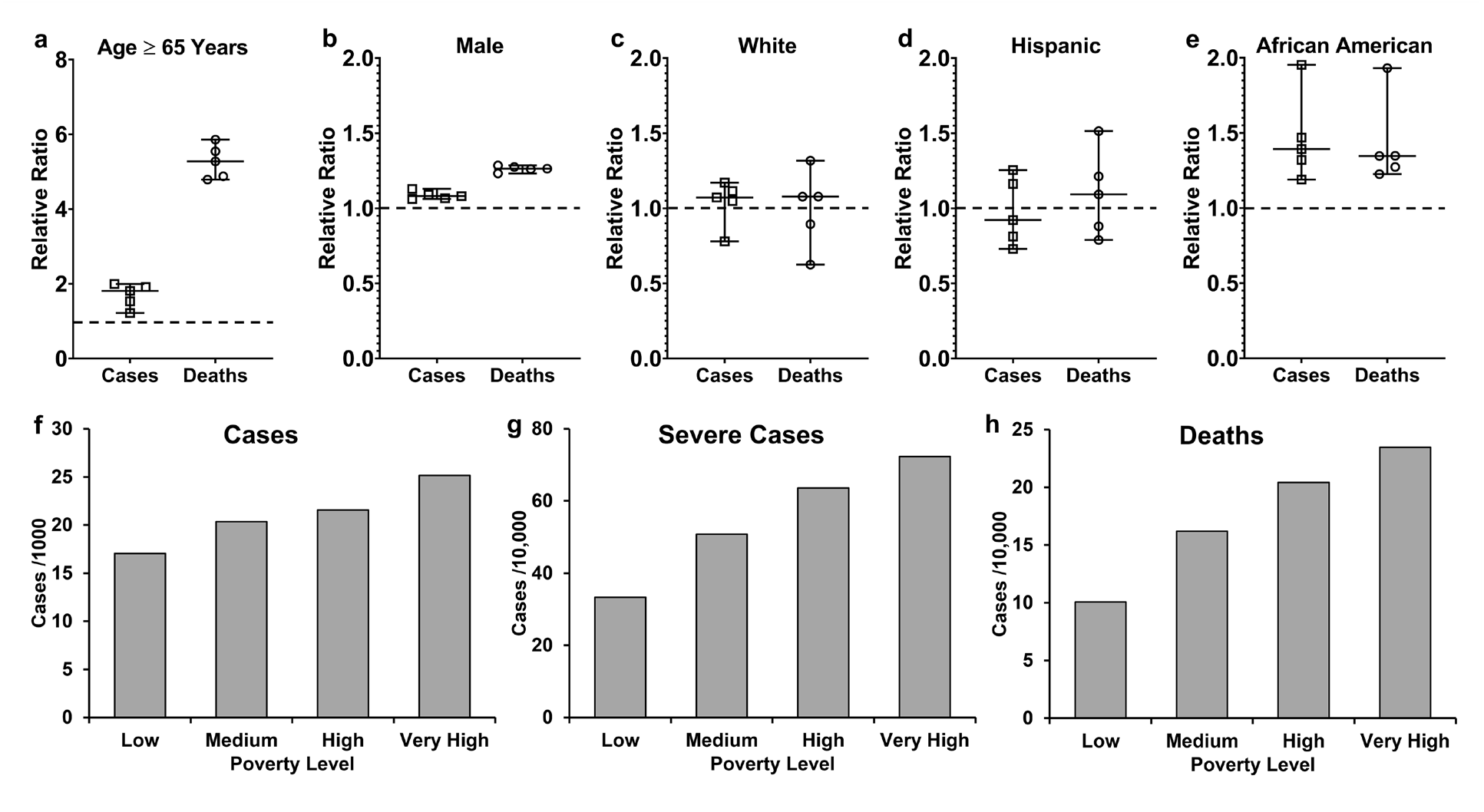
COVID-19 cases stratified according to demographic factors in New York Borough. COVID-19 cases and deaths reported by the State of New York on May 20^th^, 2020 for the counties Kings, Bronx, New York, Queens, and Richmond (NYC, 2020). (a-e) The relative ratio is the ratio of percentage of cases or death in the corresponding demographic category to the percentage county population in the same demographic category. (a) Percent population age ≥ 65 years. (b) Percent population male sex. (c) Percent population White ethnicity. (d) Percent population Hispanic ethnicity. (e) Percent population African American race. (f-h) COVID-19 cases and deaths in the New York Borough, stratified according poverty level at the zip code areas. Low <10% of residents living below the poverty line. Medium 10% to <20% of residents living below the poverty line. High 20% to <30% of residents living below the poverty line. Very high ≥30% of residents living below the poverty line. (f) Cases rates. (g) Severe case rates. (h) Mortality rates.

**Figure 8.**
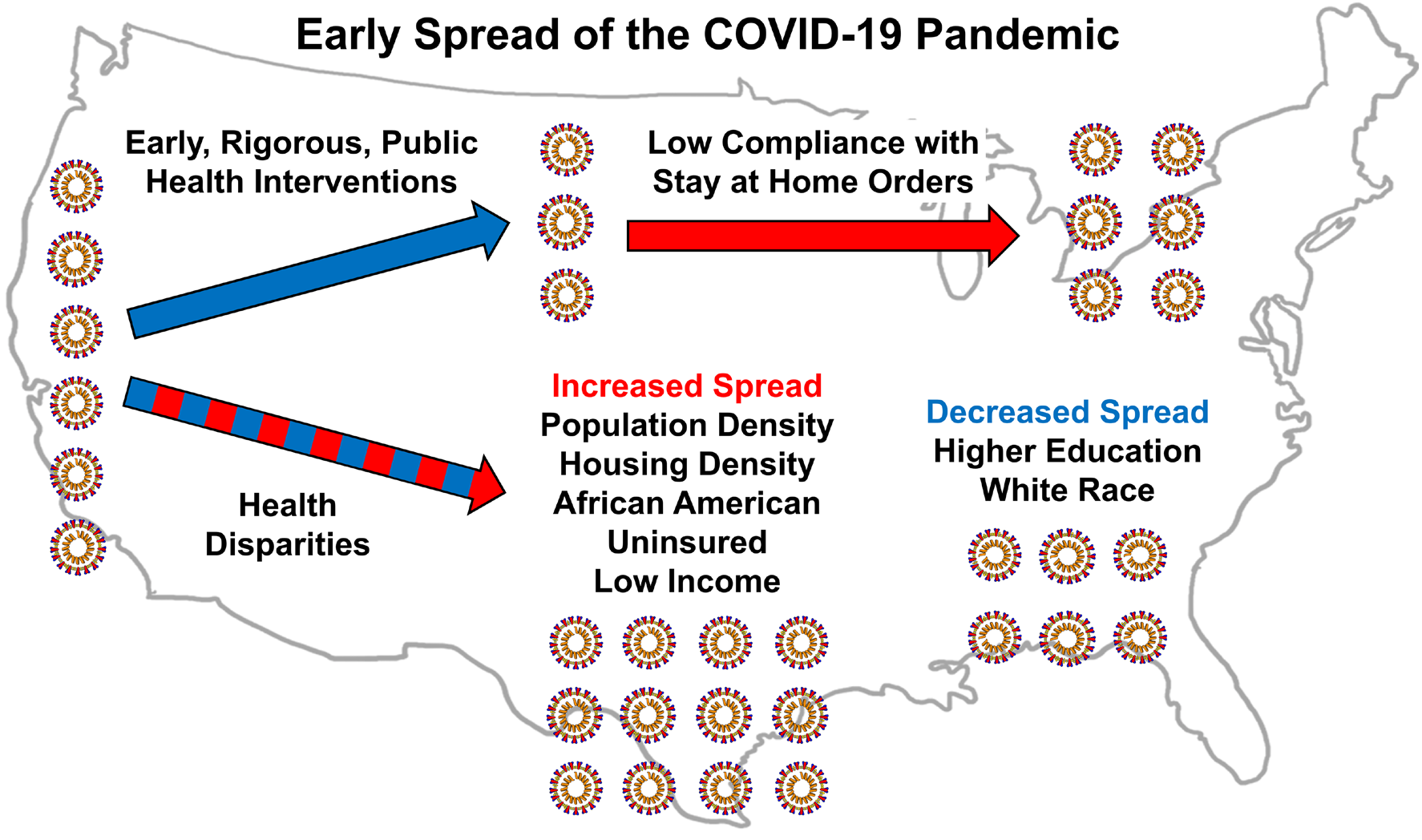
Factors that influenced the early spread of COVID-19 in the US. Three factors that affected the early dissemination of the COVID-19 pandemic in the US were public health interventions, level of compliance to stay at home orders and underlying health disparities. Early and rigorous public health interventions helped slow the spread of the Coronavirus. Low level of compliance (<50% reduction in community mobility) during the period stay at home orders were in place helped continually spread the virus. Underlying health disparities observed across the US caused disproportionate early spread of the COVID-19 pandemic.

### 3.5 Multiple regression model analyses and results

Since this study focused on 30 counties/data points, not all variables were analyzed together in multiple regression models. The response variable for the models shown was case rates for May 10^th^, 2020. In general, remarkably similar results were obtained using the other dates, mortality rates, case-fatality rates, and the slopes of lines through the case/mortality/fatality rates. Three models are reported below, one for each group of variables studied: interventions, compliance, and health disparities.

### 3.5.1 Model for public health interventions

The three explanatory variables for the intervention multiple regression model were mass gathering order duration, stay-at-home order duration, and facemask duration. All pair-wise interactions were included in the model. There were seven coefficients, including intercept, only one with a p-value < 0.05. The model statistics were F(6, 23) = 3.83, p-value = 0.0086, RSE = 71.88, R^2^ = 0.50, R^2^_adj_ = 0.37 (Supplemental Table 7 and Supplemental Figure 3). This implies that, at a single point in time, the interventions have some explanatory power in the case rates, but it is relatively weak, indicating there are other more significant factors.

#### 3.5.2 Model for compliance with stay at home orders

The six explanatory variables for the compliance multiple regression model were the % change in movement from the start of the stay at home orders to their end (or May 10^th^) for retail, grocery, parks, transit, workplace, and residential. There were seven coefficients, including intercept, none one with p-value < 0.05. The model statistics were F(6, 23) = 3.53, p-value = 0.0126, RSE = 73.33, R^2^ = 0.48, R^2^_adj_ = 0.34 (Supplemental Table 8 and Supplemental Figure 4). This implies that, at a single point in time, the compliance has some explanatory power in the case rates, but it is relatively weak, indicating that there are more significant factors.

#### 3.5.3 Model for health disparities

The ten explanatory variables for the health disparities multiple regression model were population density, housing density, % age ≥ 65 years, % male sex, % white ethnicity, % Hispanic ethnicity, % African American race, % with income below the poverty line, % with at least one year of college education, and % uninsured. Among multiple interaction assessments, one of the most effective models had 20 coefficients (including the intercept), and 13 of them had p-value < 0.05. The model statistics were F(19, 10) = 40.7, p-value = 0.0000005, RSE = 17.42, R^2^ = 0.99, R^2^_adj_ = 0.96 (Supplemental Table 9 and Supplemental Figure 5). This implies that, at a single point in time, the health disparities substantially explain the differences in case rates. When the health disparities are combined in a model with the strongest intervention and compliance variables, the intervention and compliance variables are still not statistically significant, while the health disparity variables remain significant. The strongest health disparities, which are the primary drivers of the explanatory power of the model, are population density, % Caucasian ethnicity, % African American race, % with income below the poverty line, and their interactions.

## 4. Discussion

In this study, we evaluated the disproportionate early spread of the COVID-19 pandemic in the US, with a particular emphasis on public health interventions implemented, level of compliance to those preventative interventions, and underlying health disparities across the most populous counties. We showed that by May 10^th^, 2020, the 30 study counties had vastly different case rates ranging from 7.7 to 292.2 per 10,000 (Table 1, Figure 1). Our analyses on the public health interventions showed that the early implementation of stay at home orders and face mask requirements likely resulted in lower COVID-19 cases rates for regions assessed. Better yet, when public health interventions, including restrictions on mass gatherings, stay at home orders, and face mask requirements were implemented concurrently and early, the spread of COVID-19 was significantly lower than regions that did not incorporate these measures.

While these interventions were implemented very early on in most US states with the likely exception of New York, by early July 2020, the US started experiencing continued propagation of positive COVID-19 cases that spread across many more states (Jalali et al., 2020). To help explain this phenomenon and evaluate the robustness of the nationwide stay at home orders, we evaluated the level of compliance observed at a community level. Our results showed that irrespective of the rate of spread of COVID-19, most counties displayed a low level of compliance to the stay at home orders implemented in their state. Categorical locations that displayed the lowest level of compliance with stay at home orders were parks, grocery stores, and pharmacies. The underlying level of non-compliance, in conjunction with relaxations of these public health interventions after one to two months of implementation, likely caused a suppressed level of viral spread followed by a resurgent wave. Regardless, it appears that early interventions reported herein were effective in delaying the inevitability of viral spread, thereby providing tangible evidence to support the concept of lowering the curve.

With the differences in public health responses and the varying degree of compliance to the stay at home orders observed, outcomes presented in this article cannot fully explain the vast difference we observed in the early COVID-19 case rates. However, one particular factor that is of importance to the investigators in this article is to highlight that health disparities may likely have played a large role in the disproportionate spread of COVID-19. For example, the emergence and persistence of disparities related to health often manifest through environmental, socioeconomic, or system-level factors that are complex, and may disproportionately affect minority communities (Brown et al., 2019). Differences observed between populations are closely tied to economic, social, and environmental disadvantages that may hinder a person’s ability to achieve optimal health (CDC, 2016; HP2020, 2020). In our evaluation of various health determinants across the most populous US counties, the following list of factors were all associated with amplified case rates of COVID-19 during the early stages of the pandemic. These were: population density, housing density, African American race, education, and percent uninsured.

Various social determinants of health including race and ethnicity, access to healthcare, income inequality, housing, and social support have shown to contribute to the increased COVID-19 spread and related mortality (Rollston and Galea, 2020; Turner-Musa et al., 2020; Webb Hooper et al., 2020; Yaya et al., 2020). African American individuals have also been disproportionately affected by this disease due to the presence of comorbidities associated with worse outcomes, limited COVID-19 awareness, and composing of a significant portion of essential workers in the US (Dorn et al., 2020; Holmes et al., 2020; Tai et al., 2020; Wilder, 2020). Our study findings on health disparities are similar to what has been demonstrated in previous studies. Many of the individual factors have consistently tended to cluster under broader measures such as SES. Our findings would also support the interconnectedness of the factors we found as being clustered, yet negatively compounding.

While most previous studies have focused on the underlying health disparities and their association to the spread of COVID-19, research studies have not well characterized the impact of the public health interventions on the pandemic, or the level of community compliance to these interventions. It is important to take a holistic view of multiple factors that contribute to the disproportionate spread of COVID-19 and understanding these are important for proper modeling of the viral spread and implementing targeted preventative measures (Chowkwanyun and Reed, 2020). While our investigation was limited to 30 US counties, the outcomes presented herein may serve to aid in the preparation of a framework for future investigations aimed at better serving populations at risk for greater health disparities through public health interventions.

We have identified a few limitations with this study. For example, data from the 30 most populous counties were included in this investigation. These data, unfortunately, represent a small segment of the overall population in the US, and thus it would be inappropriate to extrapolate generalizations for all regions in the US. The regions and data utilized also served as a snapshot of the early spreading of COVID-19. The investigators of this study purposefully aimed to reduce the potential introduction of a time-based bias so that public health interventions could be compared uniformly across all counties. Our results have illustrated how early COVID-19 began to spread through populous counties. Second, the mobility data we used to determine compliance to stay at home orders were derived from time-series data. We averaged the daily percent changes to give categorical data for comparison purposes. In doing so, we may likely have overlooked daily changes that may have provided a greater level of insight. However, this is beyond the scope of this study and is a potential future direction of research. Two other factors may have impacted the case rates of some of these study counties. COVID-19 testing and screening were not uniformly administered across these counties that could lead to some bias in the number of reported cases. Besides travel within a county, there is travel across counties that can likely influence the spread of COVID-19. We did not take into account this variable as it is not readily quantifiable, and is also a potential limitation of the study.

In our study, we also observed that mortality rates and case-fatality rates were higher in counties with higher case rates (Figures 2 a-d, Supplemental Figure 2 a-d). The very high case rate to mortality rate correlation (0.93) indicates a very similar disease behavior across counties. In contrast, the substantially lower-case rate to case-fatality rate correlation (0.57) indicates the presence of additional underlying factors that are causing case-fatality rates to fluctuate across counties. Whether this is due to variations in case reporting, the rapid onset of viral transmission, the overburdened medical systems, underlying health disparities, or differences in testing needs to be carefully investigated further. Our multivariate analyses showed that there are indeed other external factors that contribute to the disproportionate case-fatality rates observes across US counties. Overall, our study takes a holistic approach to characterize the early spread of COVID-19 in the US and the multiples factors that contribute to its disproportionate spread. Identifying these factors and addressing them can help mobilize the appropriate resources to implement targeted preventative measures that help promote health equality and reduce the spread of COVID-19.

## Data Availability

All datasets presented in this study are included in the article/ supplementary material.

## Author Contributions

AJ characterized the public health response, helped analyze data, and drafted the article. SK, JS, and AG assisted in analyzing data and helped with writing. BP and RG helped with writing and editing. GB helped with computational modeling of the data. JW performed statistical analyses and helped with writing and editing. TG led the study, helped analyze data, performed statistical analyses, prepared figures, and helped with writing and editing.

## Acknowledgments

We thank Dr. Jeffrey S. Wang, Infectious Disease Specialist at Kaiser Permanente, Anaheim, California, for his clinical insights.

## Supplementary Material

The Supplementary Material for this article can be found online at: (Link)

